# Interpregnancy interval and pregnancy loss in a rural South Africa: A population-based cohort study

**DOI:** 10.1101/2021.03.18.21253877

**Authors:** Y. Moodley, Kobus Herbst, F. Tanser

**Author notes:** Email address for corresponding author (Y. Moodley).

## Abstract

**Study question:** What is the relationship between interpregnancy interval (IPI) and pregnancy loss in a a rural sub-Saharan African (SSA) setting?

**Summary answer:** IPIs >60 months, but not <6 months, were associated with a higher odds of pregnancy loss in our SSA setting.

**What is known already:** IPIs >60 months are detrimental to both fetal and maternal health, while contradictory findings exist for IPIs <6 months. No studies have investigated the relationship between IPI and pregnancy loss in SSA settings, despite high pregnancy loss rates and exponential population growth in the SSA region.

**Study design, size, duration:** Population-based cohort involving 8940 women aged 16-35 years who reported two consecutive singleton pregnancies between 2000 and 2017.

**Participants/materials, setting, methods:** Study participants were from the uMkhanyakude District in KwaZulu-Natal, South Africa. We obtained data on pregnancy-related characteristics and other variables relevant to pregnancy loss from regular surveys conducted by the Africa Health Research Institute (AHRI) as part of its demographic and health surveillance platform. IPI was determined as the time in months between the end of the first pregnancy and the start of the second pregnancy. Pregnancy loss was defined as either miscarriage or stillbirth. We used an adjusted logistic regression model to investigate the relationship between IPI and pregnancy loss.

**Main results and the role of chance:** IPIs >60 months were associated with an almost three-fold higher odds of pregnancy loss (Adjusted Odds Ratio: 2.64, 95% Confidence Interval:1.71-4.09) when compared with IPIs of 6-60 months. IPIs <6 months conferred a similar odds of pregnancy loss when compared with IPIs of 6-60 months (Adjusted Odds Ratio: 0.82, 95% Confidence Interval: 0.35-1.91).

**Limitations, reasons for caution:** Possible recall bias around some of the pregnancy-related data. Inability to adjust our multivariate statistical analysis for certain sexually transmitted diseases which are known risk factors for pregnancy loss.

**Wider implications of the findings:** Family planning services in SSA should consider discouraging IPIs >60 months. Although IPIs <6 months had no impact on pregnancy loss, these should also be discouraged in SSA, given the potential socioeconomic consequences for the already vulnerable women of this region.

**Study funding/competing interest(s):** The corresponding author was supported with a postdoctoral fellowship under a National Institute of Health grant (R01 HD084233). The AHRI demographic and health surveillance platform is supported by the Wellcome Trust (201433/Z/16/Z), and the South African Population Research Infrastructure Network. No competing interests are declared.

**Trial registration number:** N/A.

## INTRODUCTION

Pregnancy loss, which includes miscarriage and stillbirth, remains an important public health problem in both developed and developing countries (Hogue, 2016). An estimated 10-20% of recognized pregnancies in developed countries end in miscarriage (Hogue, 2016), with this estimate likely to be worse for developing countries who face additional challenges relevant to women’s reproductive health (Bhasker Rao, 2000). Indeed, the disproportionate gap in pregnancy loss between developed and developing countries is clearly demonstrated in a recent analysis which found that sub-Saharan Africa (SSA) and South Asia accounted for 40.5% and 36.9% of global stillbirths between 2000 and 2015, while only 1.8% of stillbirths during the same time period occurred in developed countries (Blencowe *et al*., 2016).

Pregnancy loss has psychological consequences for the families involved (Farren *et al*., 2018), and has also been linked with subsequent maternal mortality (Reardon & Thorp, 2017). Given these implications, and the projected exponential growth of the SSA population in the next few decades (Ezeh *et al*., 2020), interest in understanding modifiable risk factors for pregnancy loss in this region has gained momentum. The interpregnancy interval (IPI), or the period of time between the end of one pregnancy and the start of the next pregnancy (Swaminathan *et al*., 2020), is perhaps one of the least understood modifiable risk factors for pregnancy loss.

A consistent finding among most published studies on this risk factor is that a longer IPI, particularly when this is >60 months, is detrimental to both fetal and maternal health outcomes (Conde-Agudelo *et al*., 2007, Kwon *et al*., 2012, Gupta *et al*., 2019). However, the data which exists for IPIs <6 months is contradictory (Conde-Agudelo *et al*., 2007, Kwon *et al*., 2012, Hanley *et al*., 2017, Gupta *et al*., 2019), and poses a challenge for family planning initiatives.

Studies which seek to clarify the relationship between IPI and pregnancy loss in SSA are rare, despite the obvious implications of this knowledge for family planning initiatives and women’s reproductive health in this region. Our study sought to address this gap in the knowledge.

## MATERIALS AND METHODS

### Study design and setting

Our study was a population-based cohort involving pregnant women from the southern region of the uMkhanyakude District in KwaZulu-Natal Province, South Africa. This region (hereafter referred to as the “study area”) has been part of the Africa Health Research Institute (AHRI) demographic and health surveillance platform for almost two decades (Tanser *et al*., 2008, Gareta *et al*., 2021). The uMkhanyakude District is characterized by high unemployment and HIV incidence rates, particularly amongst women (Welz *et al*., 2007).

### Data source

AHRI has conducted general household and individual resident surveys in the study area at least twice a year since 2000 (Tanser *et al*., 2008, Gareta *et al*., 2021). These surveys have collected the demographic characteristics, socioeconomic characteristics, and selected health-related outcomes of individuals from households in the study area. Since its inception in 2000, a total of 231179 unique individuals have participated in these surveys. There were 142079 active participants during 2018 (Tanser *et al*., 2008, Gareta *et al*., 2021). Additional detail on the information collected during the general household and individual resident surveys is provided elsewhere (Tanser *et al*., 2008, Gareta *et al*., 2021). Pregnancy notification forms are a component of the individual resident survey and are completed for each woman who reports a pregnancy between survey waves (Tanser *et al*., 2008, Chetty *et al*., 2017, Gareta *et al*., 2021). This allows for multiple pregnancies for the same woman to be documented over a period of time. The content of the pregnancy notification forms includes the woman’s maternal history prior to the reported pregnancy; date of last menses associated with the reported pregnancy; the date that the reported pregnancy ended; and the outcome of the reported pregnancy, namely singleton/multiple birth, live birth, abortion, or stillbirth (Chetty *et al*., 2017). Data from the completed pregnancy notification forms is collated in a pregnancy registry, which is updated on a regular basis.

### Study sample

We reviewed the data from the AHRI pregnancy registry to identify women aged 16-35 years old who reported having at least two consecutive singleton pregnancies between 2000 and 2017. For women who had multiple pregnancies during this time period, we included only the first and second pregnancies. Women with first or second pregnancies during the study period which ended in elective abortion were excluded from our study. Following the application of these inclusion and exclusion criteria, our study sample was comprised of 8940 women.

### Exposure

We defined the IPI as the time between the end of one pregnancy and the start of the next pregnancy (Swaminathan *et al*., 2020). This was established by reviewing the AHRI pregnancy registry and calculating the time in months between the date that the first pregnancy ended and the date of the last menses associated with the start of the second pregnancy. We categorized the IPI as follows: <6 months, 6-60 months, and >60 months.

### Outcome

Our study outcome was pregnancy loss at the second pregnancy. We adopted our definition for pregnancy loss from our prior research (Moodley *et al*.,. In press.). Briefly, pregnancy loss was a defined as a pregnancy which ended in either a miscarriage or stillbirth. We defined a miscarriage as a pregnancy which ended before a gestational age of 28 weeks was reached. This was established by reviewing the AHRI pregnancy registry data and calculating the difference in weeks between the pregnancy outcome date and the date that the same pregnancy was conceived. We defined stillbirth as delivery of a dead infant of gestational age ≥28 weeks. Stillbirth is recorded as a separate pregnancy outcome variable on the AHRI pregnancy registry.

### Statistical analyses

All statistical analyses in our study were performed using R version 3.6.2 (R Foundation for Statistical Computing, Vienna, Austria). We first performed a descriptive analysis to determine the distribution of important characteristics in our study sample. The results of our descriptive statistical analysis are presented as frequencies and percentages. We then used univariate and multivariate logistic regression models to investigate the relationship between IPI and pregnancy loss in our study sample. The multivariate regression model was adjusted for several maternal characteristics (age when pregnancy was conceived, socioeconomic status, level of education, marital status, access to healthcare services – based on distance to nearest clinic, HIV serostatus, and history of tuberculosis), as well as characteristics of prior pregnancies (number of prior pregnancies and whether the first pregnancy ended in pregnancy loss). We identified these specific covariates for inclusion in our multivariate regression model from the published literature. We obtained covariate data from the AHRI general household surveys, individual resident surveys, and the pregnancy registry. The results of our regression analyses are presented as unadjusted and adjusted odds ratios (ORs) with 95% confidence intervals (CI).

### Ethical approval

The AHRI demographic and health surveillance platform has received approval from the Biomedical Research Ethics Committee at the University of KwaZulu-Natal, South Africa.

## RESULTS

### Description of the study sample

We provide a description of our study sample, which was comprised of 8940 women, in Table I. The majority of women were between the ages of 21 and 25 years old (n=3654, 40.9%). Most of the women were in the lower socioeconomic status group (n=5366, 60.0%). Just under two-thirds of women had completed primary school (n=5816, 65.1%). Only 564 women (6.3%) were married. The vast majority of women resided within 5km of a clinic (n=8163, 91.3%). A total of 1105 women (12.4%) were HIV-positive, 2772 (31.0%) were HIV-negative, and 5063 (56.6%) did not have an HIV test done prior to conception. Tuberculosis was uncommon (n=80, 0.9%). Only a small proportion of the study sample reported having ≥4 prior pregnancies (n=389, 4.3%). A total of 178 first pregnancies (2.0%) ended in pregnancy loss. Almost three-quarters of women in our study sample conceived within 6-60 months after the first pregnancy ended (n=6689, 74.8%). A total of 135 women (1.5%) suffered pregnancy loss at the second pregnancy.

**Table I.** Description of the study sample (N=8940 women)

| Variable | n (% of N) |
| --- | --- |
| *Maternal characteristics* |  |
| **Age when pregnancy was conceived** |  |
| 16-20 years | 1428 (16.0) |
| 21-25 years | 3654 (40.9) |
| 26-30 years | 2453 (27.4) |
| 31-35 years | 1405 (15.7) |
| **Socioeconomic status** |  |
| Low | 5366 (60.0) |
| High | 3574 (40.0) |
| **Completed at least primary school** |  |
| No | 3124 (34.9) |
| Yes | 5816 (65.1) |
| **Currently married** |  |
| No | 8376 (93.7) |
| Yes | 564 (6.3) |
| **Clinic within 5km of residence** |  |
| No | 777 (8.7) |
| Yes | 8163 (91.3) |
| **HIV serostatus** |  |
| Negative | 2772 (31.0) |
| Unknown | 5063 (56.6) |
| Positive | 1105 (12.4) |
| **History of tuberculosis** |  |
| No | 8860 (99.1) |
| Yes | 80 (0.9) |
| *Pregnancy characteristics* |  |
| **Number of prior pregnancies** |  |
| 1 | 6535 (73.1) |
| 2-3 | 2016 (22.6) |
| ≥4 | 389 (4.3) |
| **\*First pregnancy ended in miscarriage/stillbirth** |  |
| No | 8762 (98.0) |
| Yes | 178 (2.0) |
| **IPI** |  |
| 6-60 months | 6689 (74.8) |
| <6 months | 445 (5.0) |
| >60 months | 1806 (20.2) |
| *Study outcome* |  |
| **Pregnancy loss** |  |
| No | 8805 (98.5) |
| Yes | 135 (1.5) |

\*Refers to the first of the two pregnancies reported for each woman during the study period.

### Statistical associations between IPI, various covariates, and pregnancy loss

We present the results of our univariate and multivariate statistical analyses in Table II. We found that the odds of pregnancy loss in women with IPIs <6 months were statistically similar to that observed for the reference group of women who had IPIs of 6-60 months (Unadjusted OR: 1.04, 95% CI: 0.45-2.39; and Adjusted OR: 0.82, 95% CI: 0.35-1.91). On the other hand, we found an IPI >60 months to be associated with an almost two-fold higher unadjusted odds of pregnancy loss (Unadjusted OR: 1.81, 95% CI: 1.25-2.62) when compared with the reference group. In our adjusted statistical analysis, an IPI >60 months was associated with an almost three-fold higher odds of subsequent pregnancy loss (Adjusted OR: 2.64, 95% CI:1.71-4.09) when compared the reference group. Our adjusted statistical analysis also found a lower odds of pregnancy loss in the 31-35 year old maternal age group (OR: 0.26, 95% CI: 0.11-0.58), while the odds of pregnancy loss were higher in mothers who had a history of tuberculosis (OR: 3.98, 95% CI: 1.34-11.80), mothers with parity of 2-3 (OR: 2.61, 95% CI: 1.76-3.88) or ≥4 (OR: 4.05, 95% CI: 1.71-9.53), and when the first pregnancy was reported to have ended in pregnancy loss (OR: 3.75, 95% CI: 1.85-7.58).

**Table II.** Results of the univariate and multivariate logistic regression analyses investigating statistical associations between IPI, various covariates, and pregnancy loss

| Variable | Unadjusted OR<br>(95% CI) | Adjusted OR<br>(95% CI) |
| --- | --- | --- |
| *Independent variable under investigation* |  |  |
| **IPI** |  |  |
| 6-60 months | 1.00 | 1.00 |
| <6 months | 1.04 (0.45-2.39) | 0.82 (0.35-1.91) |
| >60 months | 1.81 (1.25-2.62) | 2.64 (1.71-4.09) |
| *Covariates (Maternal and other pregnancy characteristics)* |  |  |
| **Age when pregnancy was conceived** |  |  |
| 16-20 years | 1.00 | 1.00 |
| 21-25 years | 1.11 (0.66-1.88) | 0.85 (0.49-1.46) |
| 26-30 years | 1.42 (0.83-2.43) | 0.65 (0.35-1.20) |
| 31-35 years | 0.85 (0.44-1.67) | 0.26 (0.11-0.58) |
| **Socioeconomic status** |  |  |
| Low | 1.00 | 1.00 |
| High | 0.97 (0.68-1.37) | 0.98 (0.66-1.45) |
| **Completed at least primary school** |  |  |
| No | 1.00 | 1.00 |
| Yes | 0.94 (0.66-1.34) | 1.14 (0.77-1.71) |
| **Currently married** |  |  |
| No | 1.00 | 1.00 |
| Yes | 1.06 (0.54-2.10) | 1.16 (0.56-2.40) |
| **Clinic within 5km of residence** |  |  |
| No | 1.00 | 1.00 |
| Yes | 2.06 (0.91-4.69) | 1.90 (0.82-4.41) |
| **HIV serostatus** |  |  |
| Negative | 1.00 | 1.00 |
| Unknown | 1.48 (0.98-2.23) | 1.44 (0.92-2.24) |
| Positive | 1.34 (0.74-2.42) | 0.99 (0.53-1.85) |
| **History of tuberculosis** |  |  |
| No | 1.00 | 1.00 |
| Yes | 3.51 (1.26-9.73) | 3.98 (1.34-11.80) |
| **Number of prior pregnancies** |  |  |
| 1 | 1.00 | 1.00 |
| 2-3 | 2.06 (1.44-2.96) | 2.61 (1.76-3.88) |
| ≥4 | 1.74 (0.83-3.62) | 4.05 (1.71-9.53) |
| **\*First pregnancy ended in miscarriage/stillbirth** |  |  |
| No | 1.00 | 1.00 |
| Yes | 3.65 (1.82-7.30) | 3.75 (1.85-7.58) |

\*Refers to the first of the two pregnancies reported for each woman during the study period.

## DISCUSSION

Our analysis reveals a harmful association between IPIs >60 months and subsequent pregnancy loss. Our study findings also suggest that IPIs <6 months do not confer a higher probability of subsequent pregnancy loss amongst SSA women when compared with IPIs of 6-60 months. We confirm the well-established association between long IPIs and adverse pregnancy outcomes. The biological mechanisms underlying this association are poorly understood. However, Zhu and colleagues have put forward two hypotheses which might explain this association. The first hypothesis is that a postpartum woman’s physiological state gradually reverts to that of a primigravid woman, which in itself confers a higher probability of adverse outcomes for a subsequent pregnancy (Zhu *et al*., 1999). The second hypothesis is that the association between IPIs >60 months and pregnancy loss is due to maternal factors which are yet to be identified (Zhu *et al*., 1999).

Long IPIs among South African women are thought to be related to early premarital fertility, followed by a gap in fertility from high levels of premarital conceptive use which enables the mother to continue her education and access employment to provide for her child, then marital fertility (Garenne *et al*., 2001). Based on our findings, family planning services in SSA should consider discouraging IPIs >60 months. Our findings for IPIs <6 months are in contrast to those from a large retrospective analysis of national health surveys conducted by Swaminathan and colleagues, which reported a slightly higher risk of stillbirth (risk difference of 0.015, 95% CI: 0.012-0.017) in SSA women with IPIs <6 months when a “between-mother” modelling approach was used (Swaminathan *et al*., 2020). It is important to note that analysis done by Swaminathan and colleagues was not adjusted for certain factors associated with pregnancy loss in SSA, such HIV and tuberculosis (Kennedy *et al*., 2012, Bekker *et al*., 2016), which we had included in our analysis. Therefore, our study findings on the relationship between short IPIs and pregnancy loss in a SSA setting are likely more accurate than that reported by Swaminathan and colleagues. Although IPIs <6 months were not associated with pregnancy loss in our study, this finding must be placed in context with the overwhelming socioeconomic challenges experienced by most SSA women. Multiple pregnancies spaced closely apart might exacerbate these challenges by reducing opportunities for SSA women to participate in empowerment activities which seek to improve socioeconomic status or educational attainment (Sonfield *et al*., 2013). It is for this reason that family planning services in SSA should also consider discouraging very short IPIs.

We also found that the odds of pregnancy loss were 74% lower in mothers aged 31-35 years old when compared with the youngest age group investigated in our study (16-20 years old). Studies from Asia and Europe suggest that the statistical relationship between maternal age and pregnancy loss resembles a U-shaped curve (Nybo Andersen *et al*., 2000, Altijani *et al*., 2018), with the trough of the curve (i.e. lowest risk of pregnancy loss) occurring between the ages of 25 and 29 years old. It is possible that the 31-35 year age group corresponds to the trough of this U-shaped curve in our SSA setting, however this hypothesis requires further investigation.

Literature from elsewhere also suggests that there is a U-shaped relationship between parity and the risk of pregnancy loss, with this risk being highest among primigravid women and women with ≥3 prior pregnancies (Gardosi *et al*., 2013). Thus, our study confirms that the risk of pregnancy loss is higher in women with several prior pregnancies, which has important implications for family planning in our SSA setting.

Tuberculosis was associated with an almost 4-fold higher odds of pregnancy loss, highlighting the importance of strengthening tuberculosis control amongst pregnant women, particularly in settings where the disease is endemic (Gounder *et al*., 2011). Lastly, our finding regarding a higher odds of pregnancy loss in mothers who had experienced pregnancy loss at the first pregnancy is unsurprising, considering that a prior history of pregnancy loss is amongst one of the most well-established risk factors for pregnancy loss (Bhattacharya & Bhattacharya, 2009, Lamont *et al*., 2015).

A major strength of our study was our use of data from a population-based cohort rather than a facility-based dataset, which increases the generalizability of our findings. Our large sample size enabled us to perform an appropriate multivariate analysis without violating the statistical rule of thumb around events per variable entered into a regression model (Peduzzi *et al*., 1996). Furthermore, use of population-based data allowed us to adjust the multivariate analysis for certain covariates which would not usually be collected during facility-based studies in our setting, such as socioeconomic status and the educational attainment of the mother. An important limitation of our study was the concern around potential recall bias for data on early miscarriages and the date of last menses. However, it is likely that the potential recall bias was mitigated by the frequency of the AHRI general household and individual resident surveys (conducted 2-3 times every year). As pregnancy loss carries a high level of stigma in lower-and-middle income countries such as South Africa, and it is also possible that some miscarriages and stillbirths were not reported by women in the study area (Goldenberg *et al*., 2011). We were also unable to adjust our multivariate analysis for sexually transmitted infections known to increase the risk of pregnancy loss, such as syphilis (Gomez *et al*., 2013), as the AHRI general household and individual resident surveys did not collect data on sexually transmitted infections besides HIV.

In conclusion, we found that an IPI >60 months, but not <6 months, was associated with a significantly higher odds of pregnancy loss in our SSA setting. We recommend that family planning services in SSA settings consider discouraging IPIs >60 months. Although we found that IPIs <6 months had no significant impact on pregnancy loss, very short IPIs should also be discouraged by family planning services in SSA settings, given the potential socioeconomic consequences for the already vulnerable women of this region.

## Data availability

The data underlying this article are available (upon reasonable request) from the AHRI Data Repository at [https://data.ahri.org/index.php/home]. Pregnancy dataset: [https://doi.org/10.23664/AHRI.RD01-04.COREDATASET.PREGNANCIES.201909], General Household Survey dataset: [https://doi.org/10.23664/AHRI.RD07-99.PIP.HSE-I.ALL.201907], HIV Surveillance dataset: [https://doi.org/10.23664/AHRI.RD07-99.PIP.HIV.ALL.202007].

## Supporting information

STROBE Checklist

## Data Availability

The data underlying this article are available (upon reasonable request) from the Africa Health Research Institutes Data Repository.

https://data.ahri.org/index.php/home

## Author’s roles

All authors contributed to the study’s conception and design, data analysis, and writing of the manuscript. All authors approved the final version of this manuscript.

## Acknowledgements

The authors are grateful to the study participants and the work and support of the fieldwork and database teams at AHRI.

